# Network Action of Subcallosal Cingulate White Matter Deep Brain Stimulation

**DOI:** 10.1101/2022.07.27.22278130

**Authors:** Vineet R. Tiruvadi, Ki Sueng Choi, Allison Waters, Liangyu Tao, Rohit Konda, Nasir Ibrahim, Otis Smart, Andrea Crowell, Patricio Riva-Posse, Robert E. Gross, Christopher Rozell, Cameron C. McIntyre, Viktor Jirsa, Robert Butera, Helen S. Mayberg

## Abstract

Deep brain stimulation (DBS) within the subcallosal cingulate cortex (SCC) alleviates symptoms of depression through an unclear therapeutic mechanism. Precise stimulation of SCC white matter (SCCwm) is thought to be necessary to achieve therapeutic response, and clinical recordings can now be used to test this hypothesis. In this paper we characterized the where, what, and how of SCCwm-DBS immediate effects, its *network action*, at therapeutic stimulation frequencies. First, using simultaneous LFP and EEG, we determined whether the effects of SCCwm-DBS are *local* at the SCC and/or *remote* at downstream cortical regions. We then charactized the spatial pattern effected by DBS across high-density EEG, finding multi-oscillatory response modes. Finally, we demonstrated that these modes are spatially consistent with white matter tracts targeted during surgical implantation. These results clarify the immediate actions of SCCwm-DBS as broad low-frequency power increases in brain regions downstream to stimulated white matter. This quantitative characterization of SCCwm-DBS network action has implications for future clinical trials, and may accelerate adaptive therapy optimization.

## 1 Introduction

Precise deep brain stimulation of the subcallosal cingulate white matter (SCCwm-DBS) alleviates symptoms of treatment resistant depression (TRD) [1, 2, 3, 4]. While early studies implemented broad DBS of the SCC region [2, 5], subsequent refinements to the SCCwm have yielded improved therapeutic responses [3, 6, 4]. This suggests a critical role for adequately stimulating the four tracts of the SCCwm in achieving antidepressant response [**?**], and early studied of evoked potentials at the SCCwm demonstrate reproducibility and specificity of the EEG response. However, it remains unclear to what extent the SCC itself is modulated by therapeutic stimulation. Clarification of the therapeutic mechanisms of SCCwm-DBS across whole-brain networks is necessary to better study and standardize therapy [7].

A first step towards understanding the therapeutic mechanism of antidepressant SCCwm-DBS is to characterize its immediate effects on whole-brain oscillations, its *network action*. Previous studies of DBS effects have implicated various neural oscillations in its immediate mechanisms, with *β* and *α* emerging consistently [8, 9, 10, 11] but recordings had limited span of the potential networks being modulated. Additionally, the focus on individual oscillations may have obfuscated However, higher-order phenomena have been reported between intracranial and electrocorticography, suggesting important interactions mediated by stimulated efferents. In the case of SCCwm, the immediate *where, what*, and *how* of its oscillatory effects remains unclear, making systematic study and improvement of the therapy challenging. This *network action* remains a critical gap in our understanding of SCCwm-DBS and efforts to improve it with adaptive strategies. Preliminary evidence of evoked responses specific to precise SCCwm-DBS show promise in verifying structural engagement [12] but electrophysiologic signatures at therapeutic stimulation frequencies associated with antidepressant response are needed. Using multimodal electrophysiology, consisting of both invasive and non-invasive recordings, we can now probe network-level effects specific to SCCwm-DBS directly in patients [13, 14]. Intracranial recordings from bidirectional DBS devices can measure oscillatory changes *local* to the brain target [15, 16, 17], while dense-array scalp EEG can measure oscillatory changes from a subset of regions *remote* to the brain target [12]. Together, these recordings can clarify the level of network engagement effected by SCCwm-DBS. Furthermore, oscillatory changes across dEEG channels can be characterized against nearby, *OffTarget*, stimulation to identify a specific spatio-oscillatory *actuation mode* across *remote* recordings. While this signal is high-dimensional, the use of machine learning and explicit computational modeling can identify any robust, conserved signals in the otherwise small patient cohort [18, 19].

**Table 1:** Patient Demographics and Data. Six patients with TRD were implanted and followed for seven months - one month without therapeutic stimulation, then the experiments done here at the onset of therapeutic stimulation, followed by six months of further study. 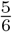 patients were responders at six months, all patients were responders at one year.

|  | Baseline HDRS17 | OnTarget Electrode | OffTarget Electrode | LFP Segments | EEG Segments |
| --- | --- | --- | --- | --- | --- |
| Pt 1 | 23.5 | L2+R1 | L1+R2 | 180 | 0 |
| Pt 2 | 20.25 | L2+R2 | L1+R1 | 180 | 0 |
| Pt 3 | 23.25 | L2+R1 | L1+R2 | 180 | 0 |
| Pt 4 | 19.25 | L2+R2 | L1+R1 | 180 | 130 |
| Pt 5 | 22.75 | L1+R1 | L2+R2 | 180 | 143 |
| Pt 6 | 20.5 | L2+R1 | L1+R2 | 180 | 103 |

Opportunistic study of neural oscillations alongside therapy is growing but comes with unique challenges due to clinical constraints. While device limitations are significant and must be taken into account, preliminarily answering mechanistic questions may yield meaningful clinical insights beyond current data-agnostic management. Experimental contrasts, combined with robust dimensionality reduction and computational links between tractography and neural recordings, is likely to provide those preliminary answers. Contrasting precise SCCwm-DBS (OnTarget) stimulation with identical stimulation away (OffTarget) we can identify actions specific to therapy. Accounting for small, heterogeneous sample sizes is more challenging, but machine learning approaches can help identify high-dimensional signals in the presence of significant, unavoidable variability. A final check to see if results across multiple measurement modalities converge, electrophysiology and tractography in particular, can increase confidence in any small-sample observation.

The primary goal of this study is to identify the recording channels and the oscillations that exhibit any measurable change immediately after initiation of SCCwm-DBS, its *network action*. Using a combination of SCC-LFP and scalp dEEG, we collected neural recordings at both *local* and *remote* scales with respect to the SCCwm target. We recorded SCC-LFP in six patients, with a subset of three having simultaneous dEEG, under both stimulation at the SCCwm target (OnTarget) and nearby (OffTarget) one month after DBS implantation, at the initiation of chronic therapeutic stimulation. To account for the small, heterogeneous cohort, we leveraged multiple convergent approaches to demonstrate oscillatory changes were robust, conserved, and supported by the targeted tractography. We first characterize at what network scale, *local* or *remote*, we see immediate oscillatory changes, before characterizing the *remote* actuation patterns specific to OnTarget DBS. First, we determined whether therapy changed oscillations at the target and/or in remote brain regions. Second, we characterized the spatial pattern of the remote response across all oscillatory bands, the *actuation mode* of the NA-SCCwm. Finally, we showed that the remote changes are consistent with brain regions downstream of the SCCwm.

## 2 Methods

### 2.1 Clinical Protocol

#### Regulatory

Six consecutive patients were implanted between June 1, 2013 and January 1, 2017 as a part of an IRB approved research protocol at Emory University studying the SCCwm-DBS for TRD (ClinicalTrials.gov Identifier NCT01984710) using inclusion and exclusion criteria described in [1]. Written informed consent was provided by each patient to participate in the study protocol (FDA IDE G130107) and the study was continuously monitored by the Emory University Department of Psychiatry and Behavioral Sciences Data and Safety Monitoring Board. All six patients were implanted with the Activa PC+S™ bidirectional DBS device, and the last three had simultaneous dense-array electroencephalography (dEEG) available (Figure 1a; See Methods 2.2). All six patients included in this study had >20 Hamilton Depression Rating Scale (HDRS17) scores over the four weeks before implantation and were clinically depressed at the time of this study’s experiments. 5/6 patients were treatment responders at 6 months, while all 6/6 were treatment responders at 12 months.

**Figure 1:**
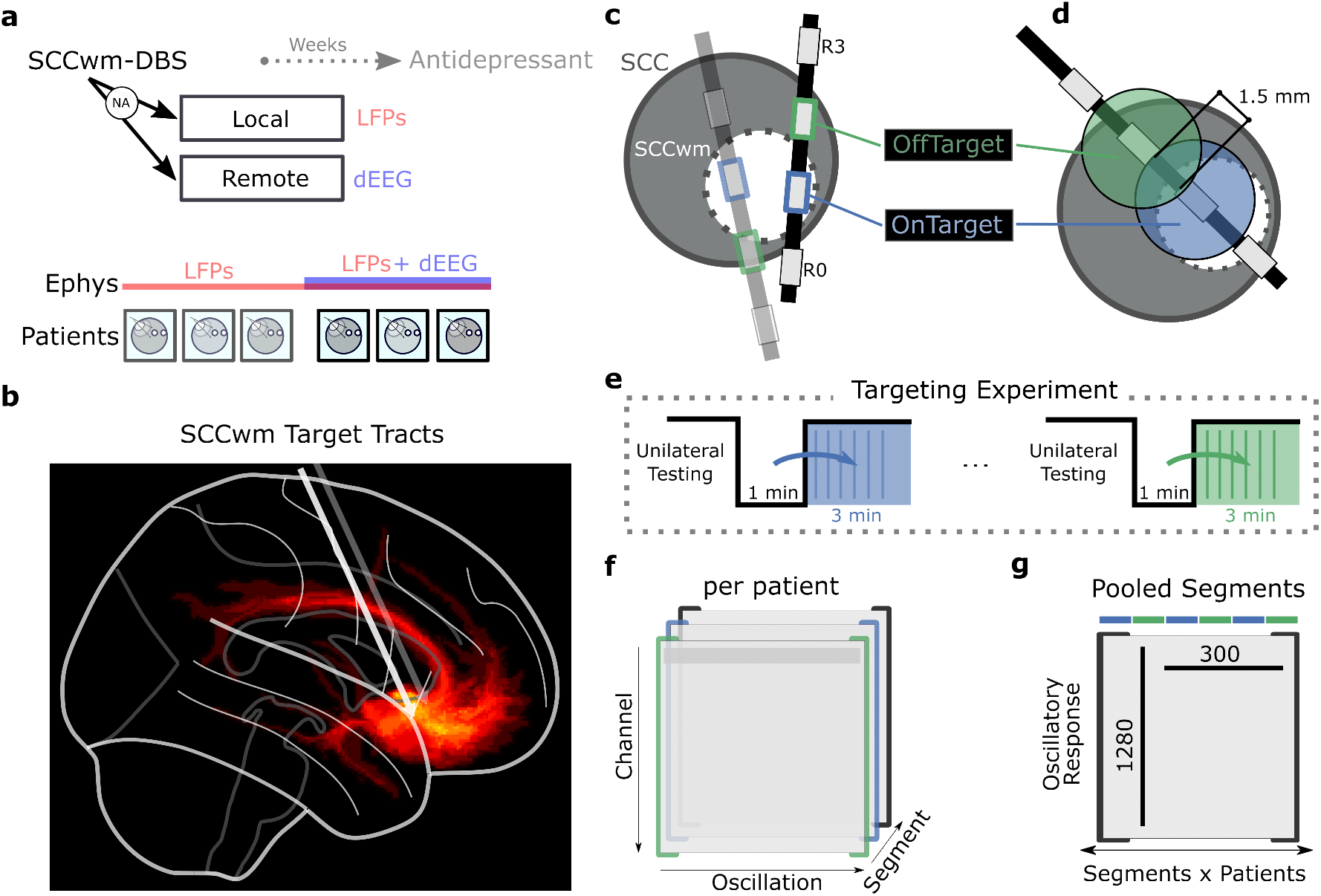
Experiment and analysis. **a**, Antidepressant SCCwm-DBS is implanted and studied over seven months. Initiation of therapy is done one month after implantation, with LFP recordings *±* EEG recordings across 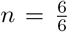 patients. **b**, The SCCwm target consists of four white matter tracts: bilateral uncinate fasciculus, bilateral cingulum bundle, forceps minor, and bilateral subcortical tracts. Two DBS leads are implanted, one in each hemisphere, after individualized connectomics-guided targeting. **c**, One of the middle electrodes from four available electrodes per DBS lead is implanted at the SCCwm target - this is the *OnTarget* electrode. The other middle electrode is either dorsal or ventral, and is the *OffTarget* electrode. **d**, OffTarget stimulation is 1.5 mm away. **e**, Targeting experiments are done at the two targets after unilateral testing (not analysed here). Each patient had two experiments, spaced over 20 min apart. Peri-stimulation recordings are normalized against the pre-stimulation baseline from the 1 min washout immediate preceding. **f**, Each patient’s recording set consists of channels x oscillatory power, for each segment of their recording in both OnTarget and OffTarget. **g**, Segments are pooled across all patients, and the features are flattened across channel x oscillatory power. The final dataset is approximately 1280 *×* 300 dimensional matrix that is analysed here.

#### Connectomic Targeting

All patients were stereotactically implanted with bilateral DBS leads (Medtronic 3387) with four 1.5 mm electrodes spaced 1.5 mm apart. DBS implantation and connectomic-guided targeting into SCCwm is described in [6]. Briefly, the target was defined by four white matter bundles within SCC region: forceps minor **Fmin**, cingulum bundle x 2**CB**, uncinate fasciculus x 2 **UF**, and subcortical fiber (Figure 1b). Personalized coordinates for the target were identified through diffusion tractography analysis of individual subject, and optimal contacts were confirmed with post-operative high resolution CT [3]. One of the middle two electrodes was implanted at the SCCwm target for chronic therapeutic stimulation, leaving another middle electrode available for an experimental contrast (OffTarget).

### 2.2 Neural Recordings

#### LFP Acquisition

The Activa PC+S™(Medtronic PLC; Minneapolis, MN, USA) recorded intracranial LFPs [16] using a differential channel from electrodes immediately adjacent to the stimulation electrode (Figure 1). LFP were sampled at 422 Hz from both left and right channels, with hardware filters set at 0.5 Hz high-pass and 100 Hz low-pass. LFP recordings were constrained to a total of 16 min at the recording parameters selected, due to limited device storage. Recording sessions lasted approximately 15 min before requiring a 20 min download process.

#### dEEG Acquisition

Dense-array EEG (dEEG) was acquired simultaneously with LFP in the last 3/6 patients using a 256-channel recording system (Electrical Geodesics, Inc., Philips) with Cz reference and sampling rate of 1000 Hz. Impedances for all scalp electrodes were maintained below 1 kΩ before all recording sessions. Patients were seated comfortably in a climate-controlled environment with head positioned on an adjustable chin rest and told to relax muscles in shoulder, neck, face.

#### Preprocessing and Segmentation

LFP recordings were assessed and corrected for nonlinear distortions secondary to impedance mismatches, a process previously described as *mismatch compression* [20]. Segmentation was done around the stimulation onset with 2 s segments.

#### dEEG Preprocessing

All dEEG recordings were bandpass filtered between 0.1 Hz to 40 Hz. ICA decomposition was performed to remove eye blinks and movement artifacts [21]. Finally, all segments were then re-referenced to the average reference with polar average reference effect (PARE) correction [22]. EEG recordings were segmented and assessed for non-stimulation artifacts (eye-blink, muscle artifacts, etc.). Segments determined to have bad channels were interpolated, unless artifacts were found across a significant number of channels and the segment was discarded. Stimulation-related artifacts are still evident however no nonlinear distortions are appreciable and baseline correction largely enables comparison of OnTarget and OffTarget responses (Figure S8).

### 2.3 Targeting Experiment

#### Targets

On each implanted DBS lead, two electrodes were stimulated in this study: OnTarget and OffTarget (Figure 1c,d). Electrode labels consist of the side (L or R) and the position number (0-3), with 0 being the furthest/most ventral electrode. OnTarget electrodes were the therapeutic electrodes specifically implanted in connectomic-guided SCCwm and used for chronic therapy. An OffTarget electrode, immediately adjacent and 1.5 mm away, was chosen on a per-patient basis to ensure a *∂*LFP channel can still be recorded simultaneously (Figure 1c) [16, 20]. The OffTarget, unlike the OnTarget, was completely unconstrained in the implantation and did not consistently engage the same structures across patients.

#### Configuration

Stimulation is experimentally delivered in three configurations at each target: unilateral left, unilateral right, and bilateral. Only the bilateral condition is analysed in this study as that is the configuration used for chronic therapy. In all bilateral stimulation initiations, left electrodes were turned on before the right, with a device-limited gap of approximately 20 s.

#### Targeting Experiment Protocol

The two different targets were delivered in two separate recording sessions spaced at least 30 min apart to allow for LFP downloads from the Active PC+S™. The full recording session lasted 15 min with three total conditions: unilateral left, unilateral right, and bilateral stimulation at a given target (Figure 1e). A 1 min washout period was done between all conditions. Only the bilateral stimulation is studied here, with the baseline being defined by the immediately preceding 1 min pre-stimulation interval. In the final three patients, simultaneous dEEG was recorded and synced using the DBS artifact.

### 2.4 Oscillatory Response

#### Frequency Domain Analysis

Power spectral densities (PSDs) are calculated for all segments using a Welch estimate with 512 FFT bins, 2 s window sizes, 0% overlap, Blackman-Harris windows yielded PSD estimates that were then log-transformed into logPSDs. Oscillatory powers are calculated by the median value in the logPSD within frequency ranges: *δ* (1 Hz to 4 Hz), *θ* (4 Hz to 8 Hz), *α* (8 Hz to 15 Hz), *β*^*\**^ (15 Hz to 20 Hz), and *γ*^1^ (35 Hz to 50 Hz). The *β*^*\**^ reported is the lower range of *β* (15 Hz to 30 Hz), adjusted for technical limitations in the PC+S™ [20]. This leads to a total feature space of 5 oscillatory band powers x 258 channels (2 LFP + 256 EEG).

#### Average Response

The baseline average oscillatory state is calculated as the feature-wise median across all pre-stimulation segments (Figure 1e) This baseline average is subtracted from all segments collected during the 3 min peri-stimulation period. The resulting *oscillatory response* vectors reflect the change from baseline at each timepoint along the first three minutes following DBS initiation.

#### Robust PCA

The Robust Principal Components Analysis (rPCA) is used to identify the largest directions of oscillatory covariance across the scalp, with rPCA chosen specifically to minimize the effects of sparse outlier channels or segments [23]. The openly available rpcaADMM implementation is used [24]. The sparsity hyperparameter is set to 0.2, reflecting approximately 20% outlier segments after visual inspection by an expert, and the resulting eigenvectors/eigenvalues are visualized as a standard PCA analysis. The major directions of variance across oscillatory features are referred to as *modes*, each mode consisting of a set of inter-oscillatory correlations and the EEG spatial pattern that those correlations are present on.

### 2.5 EEG Support Model

#### Tract Masks

Volume of tissue activated (VTA) is generated using StimVision in high resolution CT space, with stimulation parameters: 2 V to 8 V, 130Hz, and 90 microseconds [25], and the specific stimulation electrode being simulated (OnTarget or OffTarget). VTAs were then transferred to native diffusion space through high-resolution structural T1 weighted by rigid-body linear transformation and whole brain structural connectivity map was generated by probabilistic tractography in FSL [26] using a individualized bilateral VTA seeds [6]. This yields a binary voxel mask in 3D for each simulated stimulation volume, for each patient, under bilateral stimulation at both the OnTarget and OffTarget targets.

#### Average Engagement

A patient’s target-specific engagement tractography is calculated via the mean of every voxel across tract masks from all voltages 2 V to 7 V. Means are computed using NIFTI and NILearn [27] averaging across and patients in voxel-space, implemented through custom scripts. Voxels with large values are more consistent engaged across stimulation voltages than those with lower values. Average engagement tractography is calculated as the mean across patients These average tractographies are averaged across patients in each OnTarget and OffTarget condition to yield tract maps that can then be thresholded to yield a mask of voxels more likely to be engaged in each condition.

#### Network Model

The network model is taken from a 192-parcellation brain network imported from The Virtual Brain (TVB) platform [28]. Nodes that are adjacent to engaged tractography, within a threshold distance, are labeled primary nodes. Nodes that are network-neighbors to primary nodes are labeled secondary nodes, if they are not also primary nodes.

#### EEG Sensors

The position of all 256-channel dEEG sensors was imported from the system manufacturer resources. Patient-level photogrammetry was performed to align sensors to the standard map, ensuring reasonable congruence between the ideal map and the empirical map in each patient. EEG sensors that are closest to the network nodes that are, in turn, closest to the engaged tractography are labeled the *direct* channels. The EEG sensors that are closest to the neighbors of those network nodes are labeled the *indirect* channels.

#### Layer Alignment

The tractography, network model, and EEG sensor arrays are aligned visually in the same three dimensional coordinate space (Figure 2). Two thresholds are used to link tractography to network model, and network model to EEG: a distance threshold between engaged voxels and nearby network nodes, and a distance threshold between network nodes and nearby EEG channels. The coarse alignment is justified by the volume condition intrinsic to EEG recordings [29, 30] and the use of a forward-modeling paradigm that is less sensitive to small variations that can confound source modeling.

**Figure 2:**
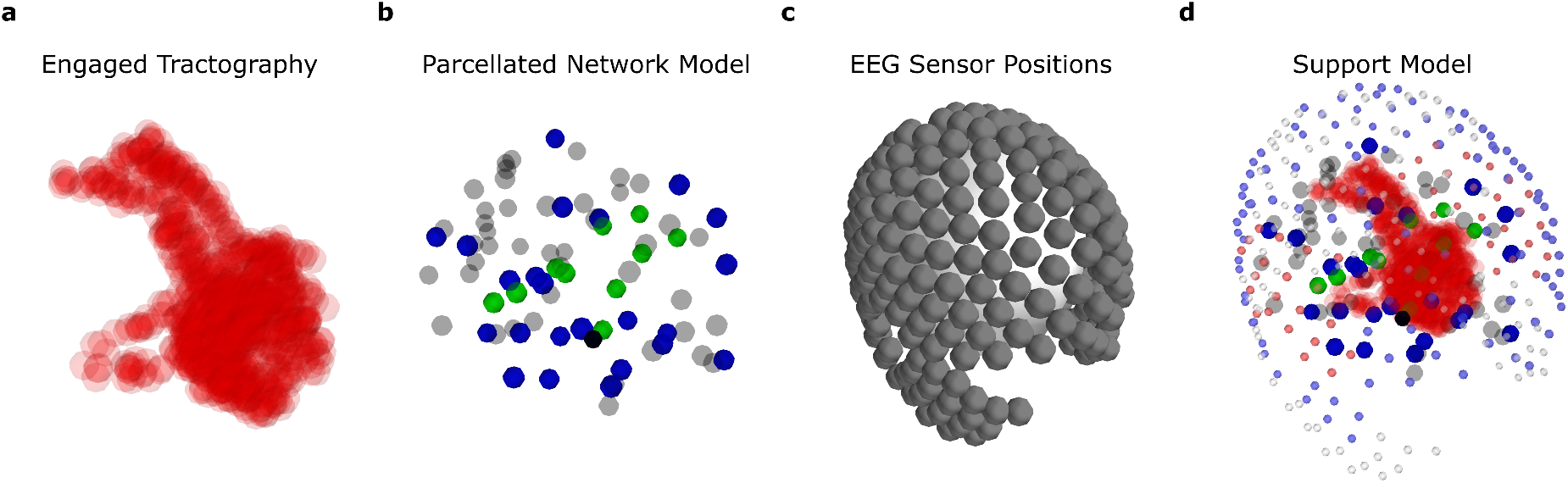
EEG Support Model. **a**, Engaged tractography is used to calculate non-zero voxels stimulated by DBS at OnTarget. Voxels are randomly sampled by a factor of 100 for 3D display. **b**, A parcellation and its network model are rendered in 3D. Nodes of the network that are closest to the tractography are identified (blue) and then used to identify their neighbors (green). **c**, All 256 dEEG sensor positions are imported from EGI Geodesic specifications. **d**, All layers are coregistered visually in the same space to yield the support model that can link engaged tractography to an EEG channel closest to the brain region it involves.

#### Statistical Test

The distributions of oscillatory changes in the direct and indirect electrodes is compared using the Mann-Whitney U-test, a nonparametric test that two samples come from the same empirical distribution [31], with *p <* 0.05 considered statistically significant.

### 2.6 Analysis Code

Analyses are implemented in Python with standard libraries, including NumPy, SciPy, and Scikit-Learn [27, 32]. Additional neuroimaging packages were also used: NiLearn and NiBabel. The custom library for model-based DBS analysis was developed independently and used here; this analysis library is available open source as a package: `DBSpace` [PyPI]. Analyses to generate the figures below are available in an open-source repository [github]. Raw data and intermediate files can be made available upon request to corresponding author.

## 3 Results

### 3.1 SCCwm-DBS effects *remote*, not *local*, action

#### LFP Response Distributions

The average (median) power in each oscillation across pooled segments is calculated for both SCC-LFP channels, and the distribution across patients is visualized (Figure 3a,b). SCC-LFP exhibited no significant oscillatory change under ONTarget stimulation (Figure3a,b blue). In contrast, OffTarget stimulation evokes SCC-LFP responses that differ significantly in *α* compared to OnTarget (*p <* 0.005, Bonferroni corrected).

**Figure 3:**
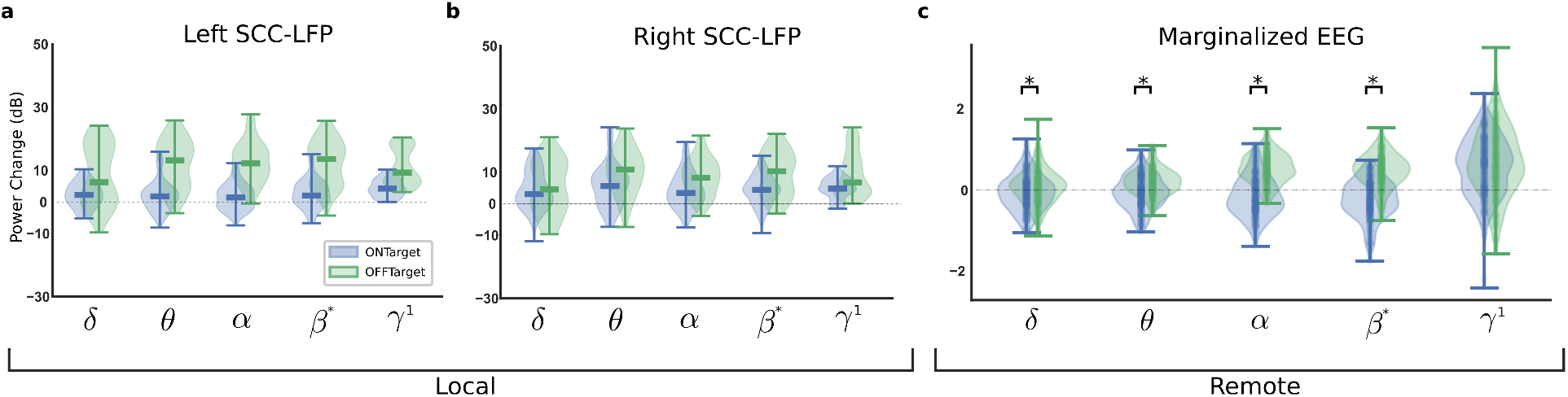
Marginal Responses in Local and Remote Recordings. LFP responses across all n=6/6 patients yield **a**, average left SCC-LFP change from baseline under ONTarget (blue) and OFFTarget (green). **b**, Right SCC-LFP oscillatory changes. Statistical significance *p <* 0.005, signified with a star (*). EEG responses in n=3/6 patients yield c) distribution of average changes marginalized across EEG channels. Statistical significance *p <* 0.005 indicated with a start (*).

#### EEG Marginalized Response Distributions

In a subset of n=3/6 patients, dEEG was recorded simultaneously with SCC-LFP under both OnTarget and OffTarget conditions. The channel-marginalized average power in each band is visualized (Figure 3c). Both OnTarget and OffTarget differ significantly from baseline, demonstrated by distributions significantly deviating from mean-zero (Figure 3c). Additionally, OnTarget and OffTarget differ significantly from each other (*p <* 0.05). *α* emerged as the most consistently present difference (Figure S9).

### 3.2 SCCwm-DBS effects patterned *remote* action

#### Individual Response Patterns

Average *α* response within each patient for OnTarget (Figure S10 top row) and OffTarget (Figure S10 bottom row) stimulation. Within each patient, OnTarget and OffTarget evoke significantly different average EEG responses (Figure 10, columns). The average (median) from pooled response segments yields the average response in each channel most consistently found in the EEG subcohort (Figure 10b).

#### Average Response Pattern

Average response patterns in *α*, calculated through feature-wise median across all available segments, demonstrates significant variability between the left and right hemisphere responses (Figure 4). Right-hemisphere *α* showed large changes under OnTarget stimulation: right parietal channels demonstrated increases while right-temporal and midline channels demonstrated decreases (Figure 4a). As a contrast, the average OffTarget response demonstrated broad *α* increases, mainly in temporal, left parietal, and right occipital channels (Figure 4b). No *α* decreases are evident in the average response.

**Figure 4:**
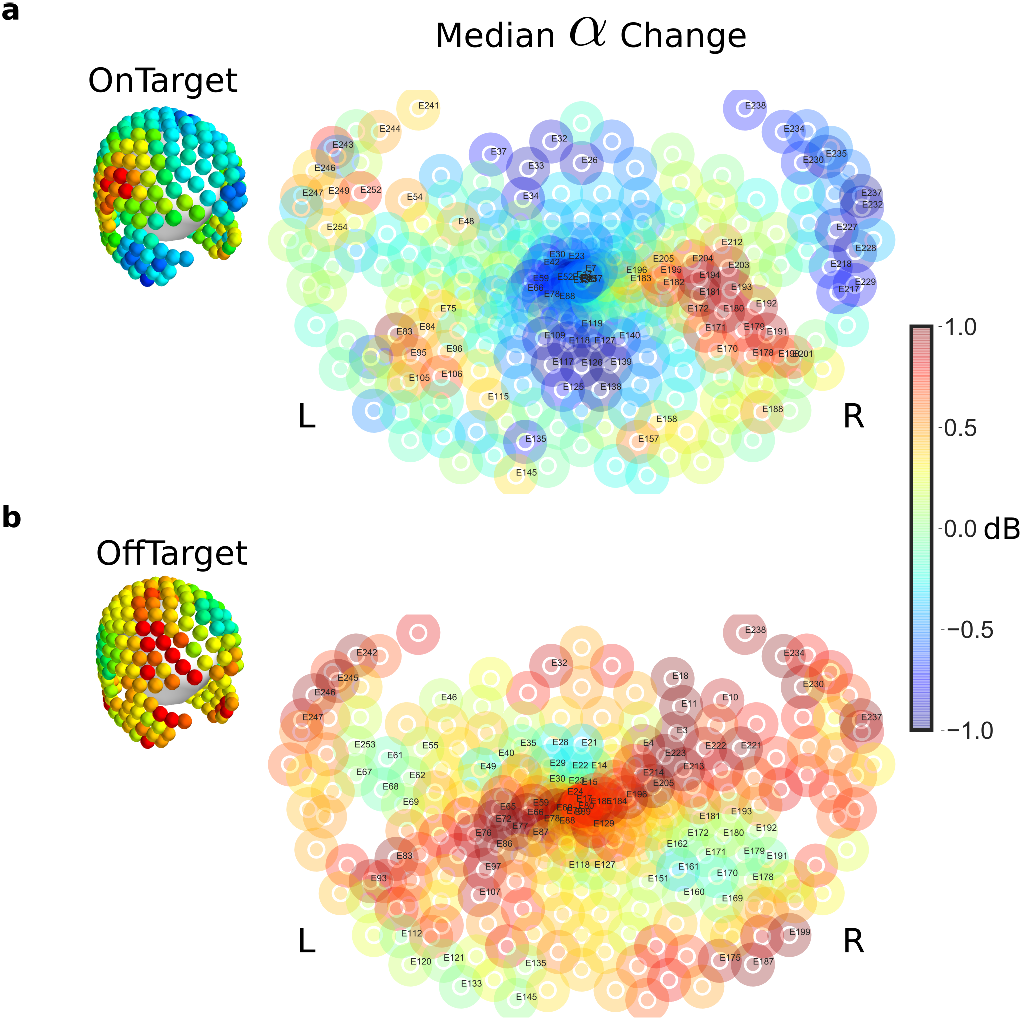
Average OnTarget *α* response pattern. Average (median) *α* change across pooled segments from all n=3/6 EEG patients. **a**, ONTarget stimulation evokes a primarily right-sided EEG change, increases in parietal channels, decreases in temporal channels. **b**, OFFTarget stimulation evokes increases in *α* across many channels bilaterally.

### 3.3 Network Action Modes

#### Patterned Variance

Robust dimensionality reduction across all features (oscillations x channel) and segments demonstrated two major directions of variance (Figure 5a). The first mode (PC0) accounted for 91% of the variance, while the second mode (PC1) accounted for 8%, with remaining modes negligible. Each mode consists of an oscillatory correlation pattern, and the spatial distribution that those patterns are present (Figure 5b).

**Figure 5:**
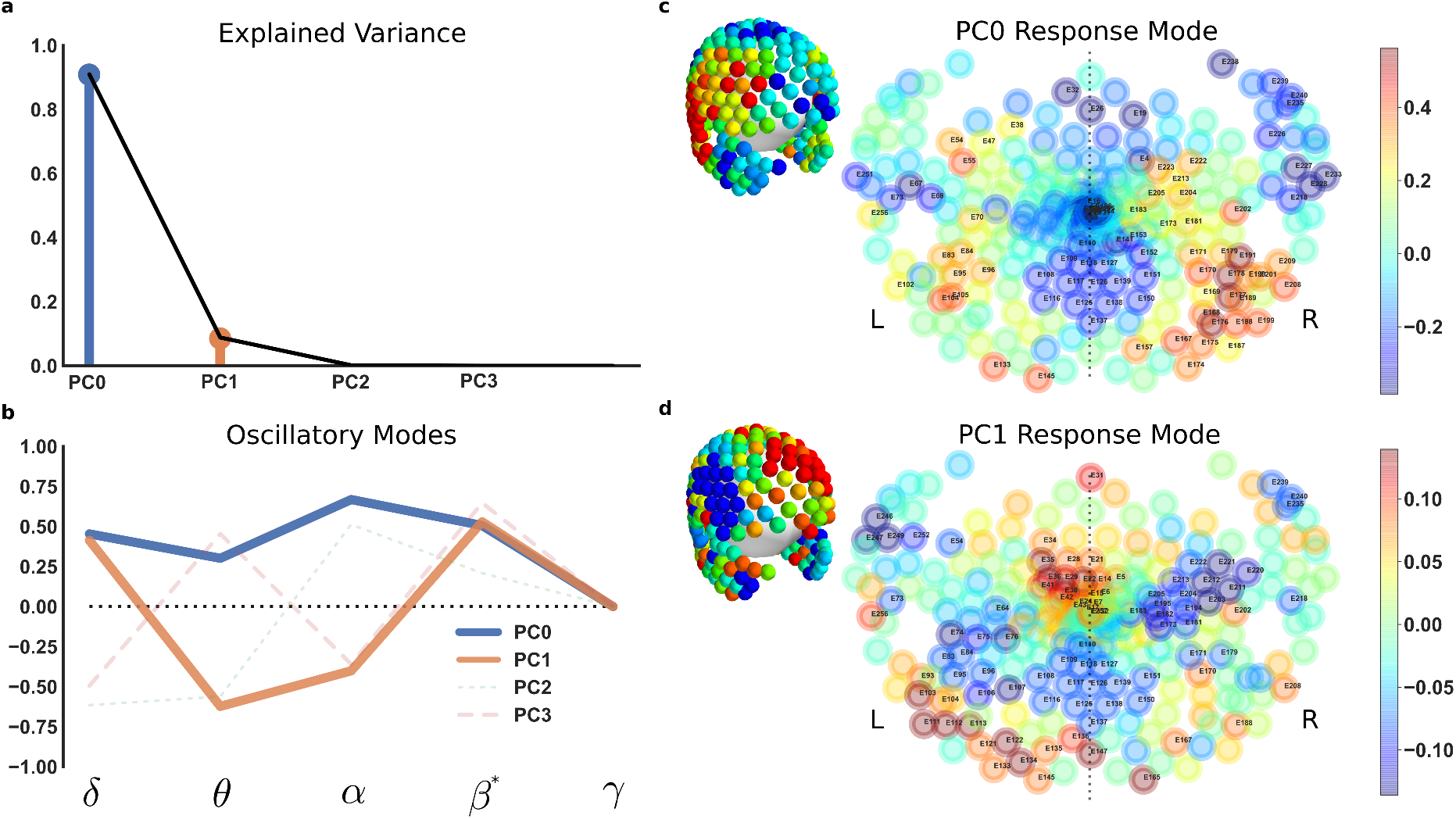
**OnTarget Actuation Modes** measured in EEG. **a**, Robust PCA identified two major directions of variance, or *modes*, in oscillatory power observed across all segments. **b**, The two modes consist of (1) all low-frequencies correlated (blue) and (2) *δ* + *β* anticorrelated with *θ* + *α* (orange). **c**, The first mode exhibits right hemisphere dominance, with positive coefficients in right parietal channels, and negative coefficients in right temporal and midline channels. **d**, The second mode exhibits a more symmetric response, with positive coefficients at midline over the SCC and negative coefficients across bilateral parietal and occipital channels.

#### Primary Modes

The primary mode consisted of all low-frequency oscillations correlated (Figure 5b PC0). The spatial distribution of this correlation was asymmetrically present on right and midline (Figure 5c). In particular, right parietal channels exhibited increases in power across all low-frequencies, while right-temporal and midline channels exhibited decreases. Frontal channels exhibited decreases in this mode as well, with smaller magnitude than the posterior midline and right temporal channels.

#### Secondary Modes

The secondary mode consisted of *δ - β*^*\**^ anticorrelated with *θ - α* (Figure 5b PC1). The spatial distribution of this correlation was more symmetric, with increases at the apex channels, and decreases in parietal channels. Scattered increases were seen in left posterior channels.

### 3.4 Modes Align with Tractography

#### Target Engagement

Target engagement is calculated under OnTarget and OffTarget stimulation for all patients, averaged, then threshold masked (Figure S11). Within the EEG subcohort, we visualize where the average voxel value is larger in OnTarget than OffTarget (Figure 6b) and vice versa (Figure 6b). OnTarget preferentially engages right-CB over left-CB, and does not appreciably engage left-UF (Figure 6a). In contrast, OffTarget engages uncinate fasciculus, more thoroughly in the left than the right (Figure 6b). Both targets engage FM symmetrically along the axial axis, and engage some voxels between hemispheres.

**Figure 6:**
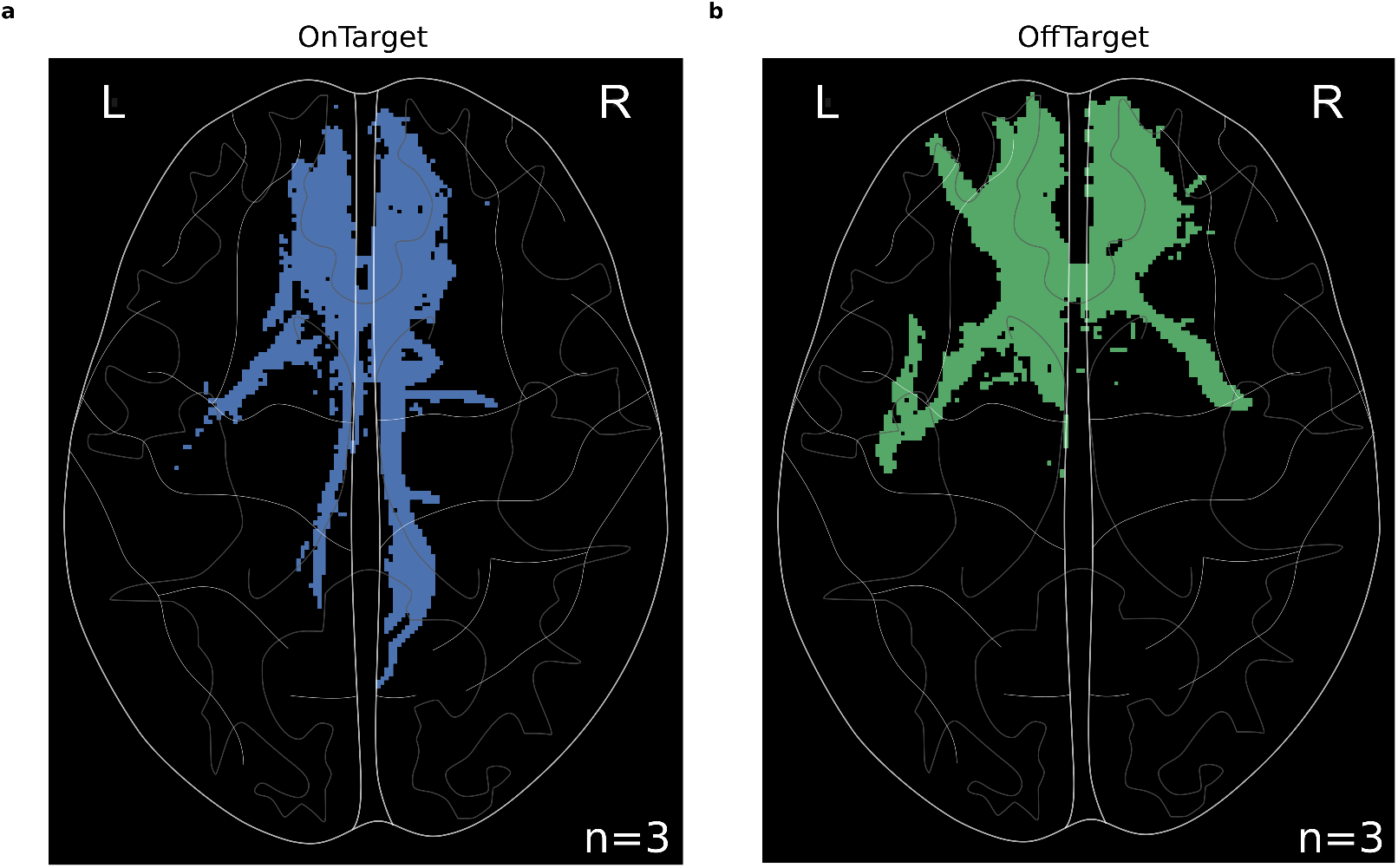
**Engaged tractography at targets** in EEG subcohort. **a**, Axial mask of above-threshold voxels in OnTarget vs OffTarget. **b**, Axial mask of above-threshold voxels larger in OffTarget than OnTarget.

#### Direct and Indirect Channels

The support model calculated for Patient 4 (Figure 7a) was used to generate two EEG masks: a direct mask and an indirect mask (Figure 7b,c). This mask was applied to the patient-specific OnTarget response, and the resulting *α* power changes in the direct and indirect masks were plotted as histograms (Figure 7d). Direct effects see an increase in *α* while indirect effects see a *α* decrease (*p <* 0.05 Kolmogorov-Smirnov Test).

**Figure 7:**
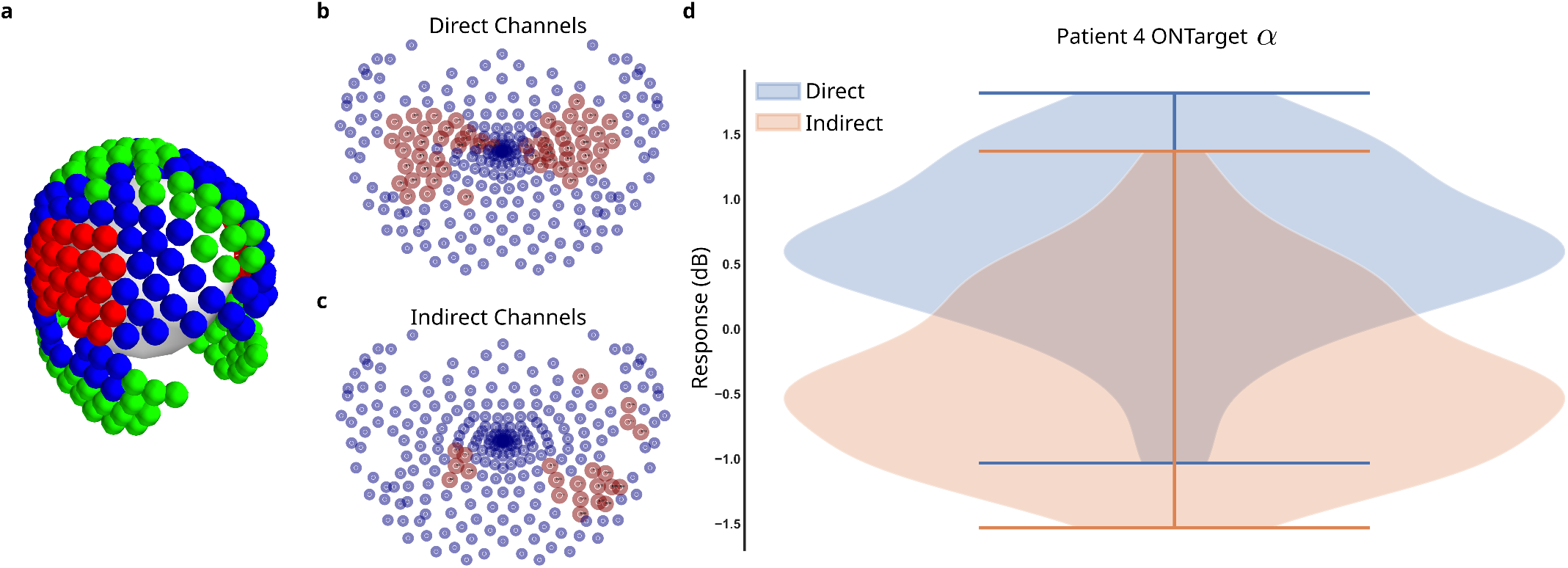
Direct and Indirect Effects in EEG. **a**, An EEG support model is constructed to identify the EEG channels closest to directly engaged brain network regions (red) and closest to indirectly engaged brain network regions (blue). **b**, EEG channel masks for direct channels demonstrate positive coefficients in bilateral parietal channels. **c**, Indirect channel mask demonstrate positive coefficients in right **d**, The empirical response from Patient 4 is masked with direct and indirect to generate response distributions in *α*. Direct channels demonstrate *α* increases while indirect channels demonstrate *α* decreases (Mann-Whitney U-test *p <* 0.05).

## 4 Discussion

In this study we used a novel set of LFP and dEEG to characterize the immediate, specific effects of precise SCCwm-DBS at therapeutic stimulation frequencies. This *network action* demonstrated effect predominantly *remote* to the SCC target, consistent with cortical regions downstream of stimulated white matter. Immediate oscillatory changes in these regions exhibited inter-oscillatory dependencies, with two major modes of effects, that aligned with the stimulated white matter tracts. These preliminary observations clarify outstanding questions in antidepressant DBS within the SCC, and our demonstration aligns empirical electrophysiology with tractography to yield a preliminary characterization of SCCwm-DBS network action at therapeutic frequencies.

### 4.1 SCCwm-DBS effects no locally measurable change

Oscillatory changes in whole-brain electrophysiology can reflect direct mechanistic effects of DBS and recent advances in clinical DBS hardware enable simultaneous deep brain and surface level recordings alongside active stimulation. Using combined SCC-LFP and scalp dEEG, we found that OnTarget stimulatio of the SCCwm therapeutic target evokes no changes *locally* at the SCC target (Figure 3a). Perhaps surprisingly, OffTarget stimulation inconsistently evoked large oscillatory changes (Figure 3b) demonstrating that *local* engagement was measurable, but not present in any OnTarget stimulations. While it is possible that the *∂*LFP recordings of the PC+S miss a large signal shared between recording electrodes, the observation of OffTarget conditions evoking large changes makes this unlikely. Additionally, while the PC+S™ has well described artifacts [16, 33, 34] and the presence of mismatch compression can distort oscillatory recordings [20], those artifacts would bias observations towards seeing spurious increases. Together, these observations strongly support the hypothesis that the SCC is not directly modulated by therapeutic SCCwm-DBS, and that targeting the SCC is not consistent with testing therapy.

### 4.2 Remote responses are patterned in *α*

In contrast with the SCC-LFP, EEG demonstrated significant changes under OnTarget stimulation, demonstrated in the channel-marginalized response (n=3/6, Figure 3c). The channel marginalization is done across the median oscillatory power change across all segments, reflecting an ensemble change and demonstrating the unlikelihood that OnTarget and OffTarget are sampled from the same distribution (the null). Marginalized across channel, the *α* and *β*^*\**^ oscillatory powers demonstrate significant, consistent deviations from zero (Figure 3c) indicating that different patterns of EEG changes are evoked across the scalp. While DBS has historically focused on *β* oscillations[10, 35, 36], including in our own group’s intraoperative observations [13, 37], *α* has been shown to be modulated by neuromodulation [8, 38] and to be linked to depressed mood [39, 40, 41, 42, 43].

To analyse these patterns directly, the pre-marginalized *α* changes are plotted for each channel (Figure 4). We saw OnTarget and OffTarget evoke visually distinct patterns across the measurable scalp (Figure 4), supporting the specificity of precise SCCwm-DBS compared to nearby stimulation. The OnTarget response exhibits an asymmetry in *α*, with right parietal increases and right temporal decreases, along with midline decreases. This median response reflects channels that exhibit any significant change at any point during the first three minutes of stimulation, with the final patterns not necessarily reflecting any instantaneous EEG state.

### 4.3 Network Action Modes

Oscillations do not change independently and characterization of coordinated changes is necessary to fully capture effects of DBS. We observed coordinated changes within two distinct modes effected by SCCwm-DBS (Figure 5). The first mode, with *α* as the largest component, evokes a right-hemisphere dominant pattern (Figure 5b,c). The correlation amongst all oscillations suggests large-scale, synchronized depolarizations, though more detailed investigation of the dynamics immediately following stimulation onset is needed [44]. The second mode, with *δ - β* anticorrelated with *θ - α*, is also present and suggests a secondary response more symmetrically mediated by brain connectivity, not axonal modulation. The symmetry of this second mode suggests a more balanced, secondary response not mediated by the engaged white matter that demonstrates significant asymmetry (Figure 5d). Notably, the oscillatory correlations in this second mode are aligned with a chronic depression readout identified in the SCC [45]. This mode’s positive coefficients in channels at the middle of the midline are consistent with the spatial extent of bilateral SCC, though it more direct links between short- and long-term activity using EEG is needed to interpret more completely.

### 4.4 Actions Along Tracts

In all patients, OnTarget consistently engages four white matter tracts: cingulum bundle, forceps minor, uncinate fasciculus, and subcortical fibers [3, 7]. Changes in the EEG under OnTarget stimulation were seen mostly in the right hemisphere, with *α* observed to have the largest change. Unlike OnTarget, OffTarget stimulation responses did not exhibit *α* decreases, potentially because of the significant variability between patients in the tracts engaged by unconstrained OffTarget stimulation (Figure 4). This provides a hint that the indirect effects of SCCwm-DBS on brain regions downstream of the stimulated target involve an *α* decrease that is not coherent enough to observe in group-level OffTarget response. While source-localization approaches are being increasingly used to study DBS, a forward modeling approach from tractography to EEG channels may provide a more robust alternative in the setting of electrically active electronics. Combined with a brain network model, the forward modeling approach may be able to tease apart both direct and indirect effects of therapeutic stimulation.

To directly test this hypothesis, we developed a *support model* approach to link electrophysiology and engaged tractography through a network model. In a single patient, we see that EEG sensors closest to direct nodes demonstrated *α* increases, and EEG sensors closest to downstream indirect nodes demonstrated *α* decreases (Figure 7). *α* power has been shown to be inversely proportional to the level of synaptic input into a brain region [46] and reflective of large scale disconnection between brain regions [47]. Our demonstration of the engagement of right-CB corroborates recent results implicating right-CB specifically as a target that must be optimally stimulated to achieve more rapid therapeutic response [4, 48]. *α*, in particular, admits a parsimonious explanation of DBS as a blockade of synaptic inputs that results in increased downstream idling or rerouted processing towards more environmentally salient inputs. The asymmetric modulation of right hemisphere *α* by SCCwm-DBS may correct intrinsic imbalances, though the ability to link our results with prior literature is challenged by our broader coverage of the brain.

### 4.5 Limitations

A major limitation in this study is the cohort size of n=6 total, with only n=3/6 having both LFP and EEG, limiting the generalizability of the work to broader DBS. ML can identify and assess high-dimensional patterns in small datasets, but limitations in generalization must be addressed by increased sample sizes. [49, 50]. Second, we only measure three minutes after stimulation onset due to limitations in the device storage, and we average through those three minutes, diluting away transient dynamics that we’ve reported elsewhere [14]. This likely ignores rich dynamics inside the measured time window and certainly misses slower effects of DBS, like neurotransmitter depletion and plasticity [51], Relatedly, the bilateral stimulation analysed in this study occurred at the end of unilateral experiments, potentially confounding our results with priming effects if the 1 min washout period between conditions was insufficient. Third, due to device limitations in the PC+S™, we did not analyse *γ* oscillations, potentially missing critical effects of stimulation. Fourth, the stimulation frequency is therapeutic but the amplitude was constant-current 6 mA while therapeutic parameters were set at constant-voltage 3 V to 5 V [52, 53, 54, 55], which may reflect suboptimal over-engagement of cingulum bundle [4]. The tract engagement was calculated across a range of voltages (Methods 2) to mitigate this, but further work is needed to compare network action as a function of stimulation amplitude. Finally, the lack of source-localization in this work prevents direct inference of the brain regions driving observed EEG changes. The support model approach developed here is a forward-modeling approach and likely less sensitive to small variations in conductance caused by the DBS device that can confound inverse modeling efforts, like source-localization[56, 57].

### 4.6 Comments on Mechanism

DBS guided by connectomics is emerging as a critical tool in standardizing, studying, and improving DBS across various disorders and indications [58] and electrophysiologic correlates will be critical to improving targeting In the case of antidepressant SCCwm-DBS, we see that precise stimulation evokes a distinct brain state in brain regions aligned with engaged tractography. Notably, this engagement is asymmetric in *α*, potentially linking our empirical observation to a long-standing hypothesis of *α* asymmetry in MDD [43]. Importantly, this oscillatory response is measurable entirely with noninvasive EEG, providing the first broadly usable signature of SCCwm target engagement at therapeutic stimulation frequencies, though further work is needed to demonstrate generalizability and clinical utility.

In parallel with clinically useful measures, direct study of DBS effected dynamics is growing and early evidence supports the idea that DBS influences symptoms via modulation of pathologically perturbed dynamics in brain networks[59, 60]. With respect to the cell-level targets, evidence consistently supports an axonal modulation at therapeutic parameters [61, 51, 62]. Here, we clarify the network-level mechanism and find evidence supportive of blockade of signaling along right cingulum bundle over the initial treatment initiation. More effort is needed to understand the *dynamics* that SCCwm-DBS evokes across scalp measurements [14] and the link to long-term *δ - β* changes that track with depression symptom recovery [45]. The support model approach taken here can be extended with neural mass models to more directly link tractography, dynamic responses, and candidate DBS mechanisms for likelihood-based inference of long-term therapeutic mechanism.

## 5 Conclusion

SCCwm-DBS at therapeutic stimulation frequencies effects a *remote*, not local, network action involving multiple oscillatory bands. This remote action is predominantly in the right hemisphere during bilateral stimulation, and is specific to precise SCCwm-DBS. The action consistent of two major spatio-oscillatory modes: a primary broad low-frequency correlation, and a secondary *δ* + *β* anticorrelated with *θ* + *α*, consistent with chronic SCC activity that reflects recovery from depression [45]. Further work is needed to verify this network action in larger cohorts with consistent neural recordings, but this preliminary characterization clarifies the immediate actions of antidepressant DBS targeted at the SCC, with implications for future clinical trials and therapeutic optimization. Future efforts should focus on developing machine learning classifiers and dynamical systems models of precise SCCwm-DBS across disease indications.

## Data Availability

Intermediate data can be made available upon request to corresponding author.

https://github.com/virati/cortical_signatures

## Acknowledgment

Sinead Quinn and Lydia Denison for assistance in patient coordination. Ashan Veerakumar for assistance in data collection. Dolu Obatusin for assistance in code and data repository setup.

## Funding

Funding support was provided by the Whitaker International Foundation, National Institutes of Health (UH3NS103550, R01MH106173), Hope for Depression Research Foundation and European Union’s Horizon 2020 Framework Programme for Research and Innovation under the Specific Grant Agreement No. 945539 (Human Brain Project SGA3). Implanted devices used in the work were donated by Medtronic, Inc. (Minneapolis, MN).

## Conflicts of Interest

CCM is a paid consultant for Boston Scientific Neuromodulation, receives royalties from Hologram Consultants, Neuros Medical, Qr8 Health, and is a shareholder in the following companies: Hologram Consultants, Surgical Information Sciences, CereGate, Autonomic Technologies, Cardionomic, Enspire DBS. HM has a consulting agreement with Abbott Labs (previously St Jude Medical, Neuromodulation), which has licensed her intellectual property to develop SCC DBS for the treatment of severe depression (US 2005/0033379A1). RG serves as a consultant to and receives research support from Medtronic, and serves as a consultant to Abbott Labs. The terms of these arrangements have been approved by Emory University, Icahn School of Medicine, and Case Western Reserve in accordance with policies to manage conflict of interest. All other authors have no COI to declare.

## 6 Supplementary*

**Figure 8:**
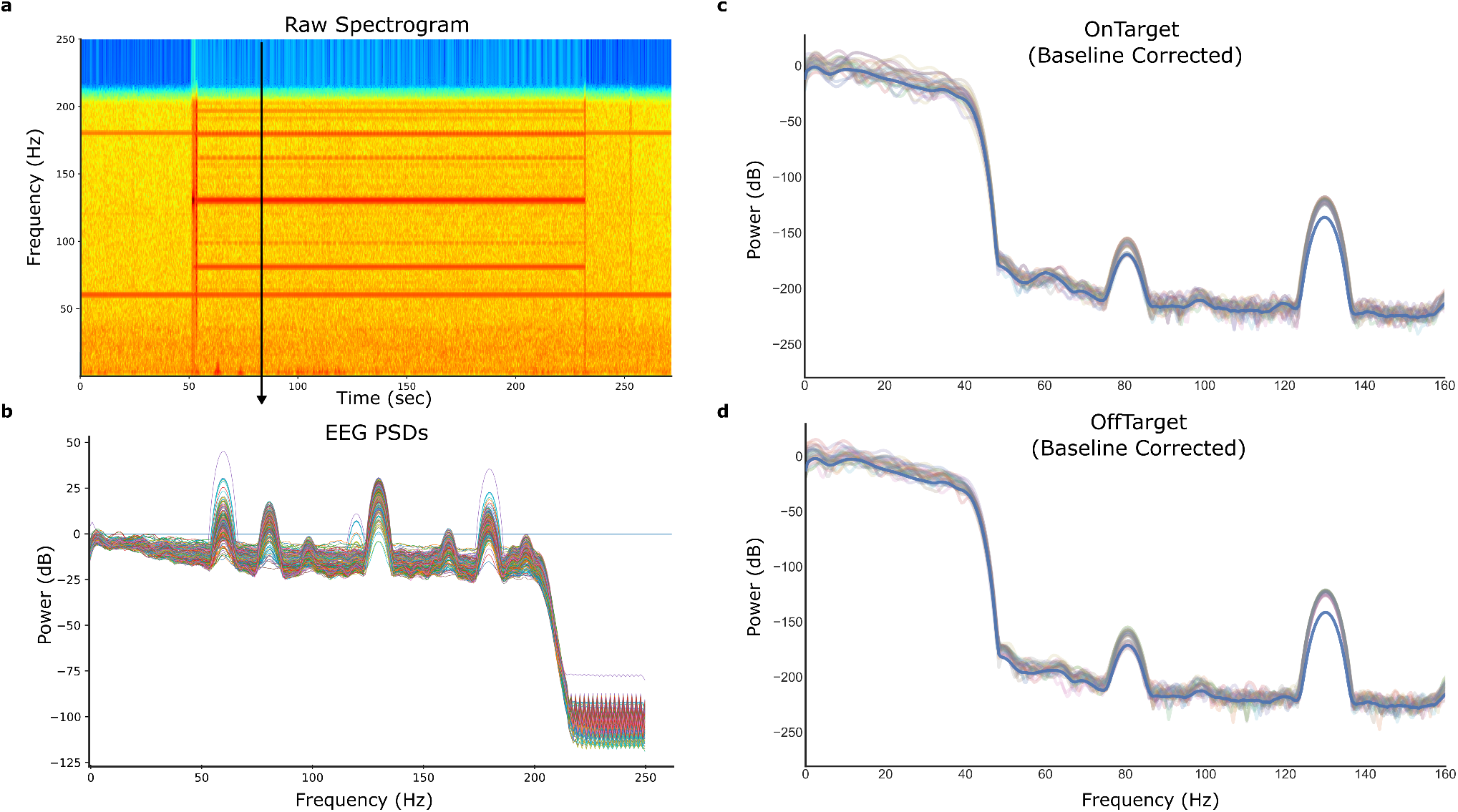
EEG artifacts are linearly separable. **a**, Spectrogram of EEG recordings demonstrates artifacts without concerning signs of nonlinear gain compression. **b**, The PSDs for all EEG channels in the same recording. The shape of the PSD and the resulting artifacts are linearly separable as a part of standard frequency-domain analyses. **c**, OnTarget PSDs after baseline correction and filtering below 50 Hz demonstrates clean oscillatory activity. **d**, OffTarget PSDs are similarly clean.

**Figure 9:**
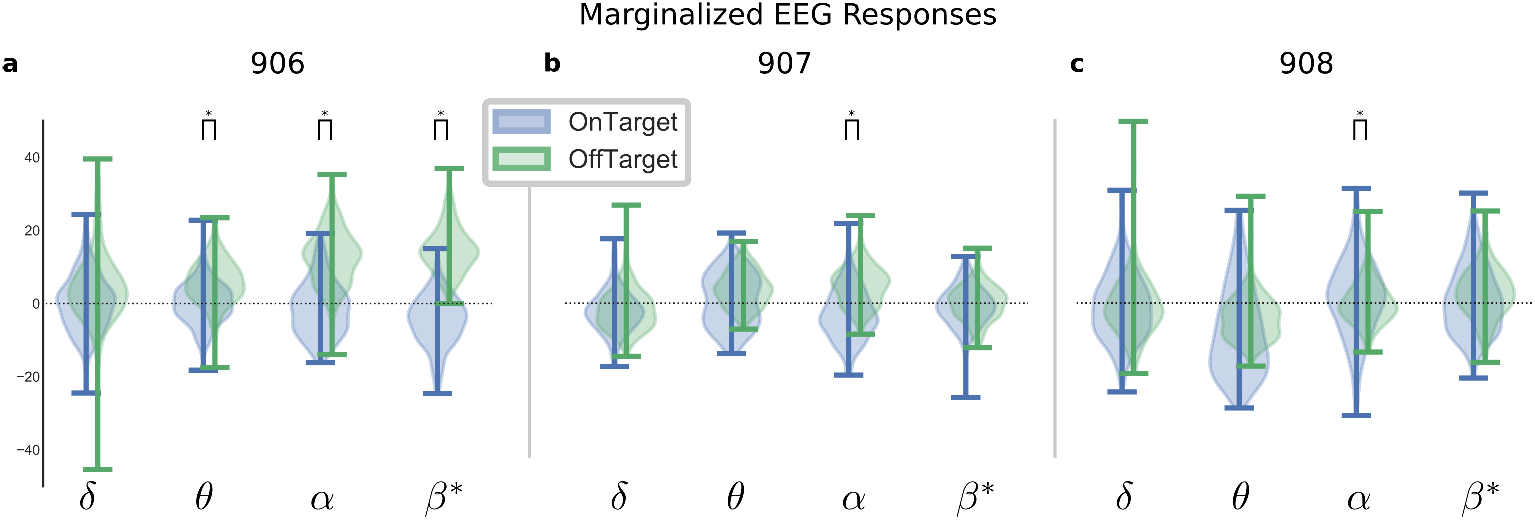
Per-patient EEG response distributions. **a**, Patient 4, **b**, Patient 5, **c**, Patient 6 distributions of average EEG response across channels.

**Figure 10:**
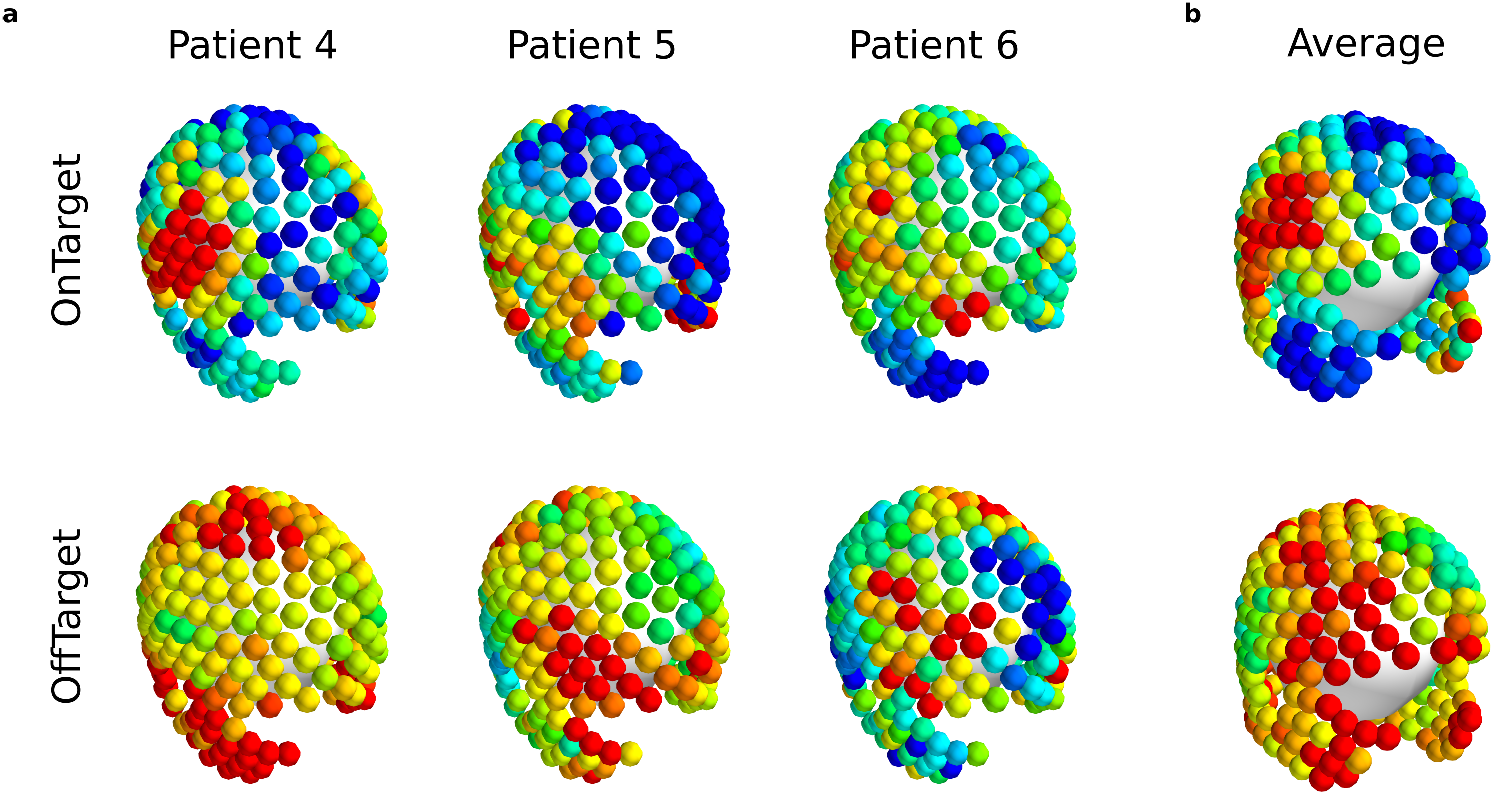
*α* response patterns per-patient. **a**, Average *α* across OnTarget (top row) and OffTarget (bottom row) stimulation in each patient **b**, with the median across pooled segments.

**Figure 11:**
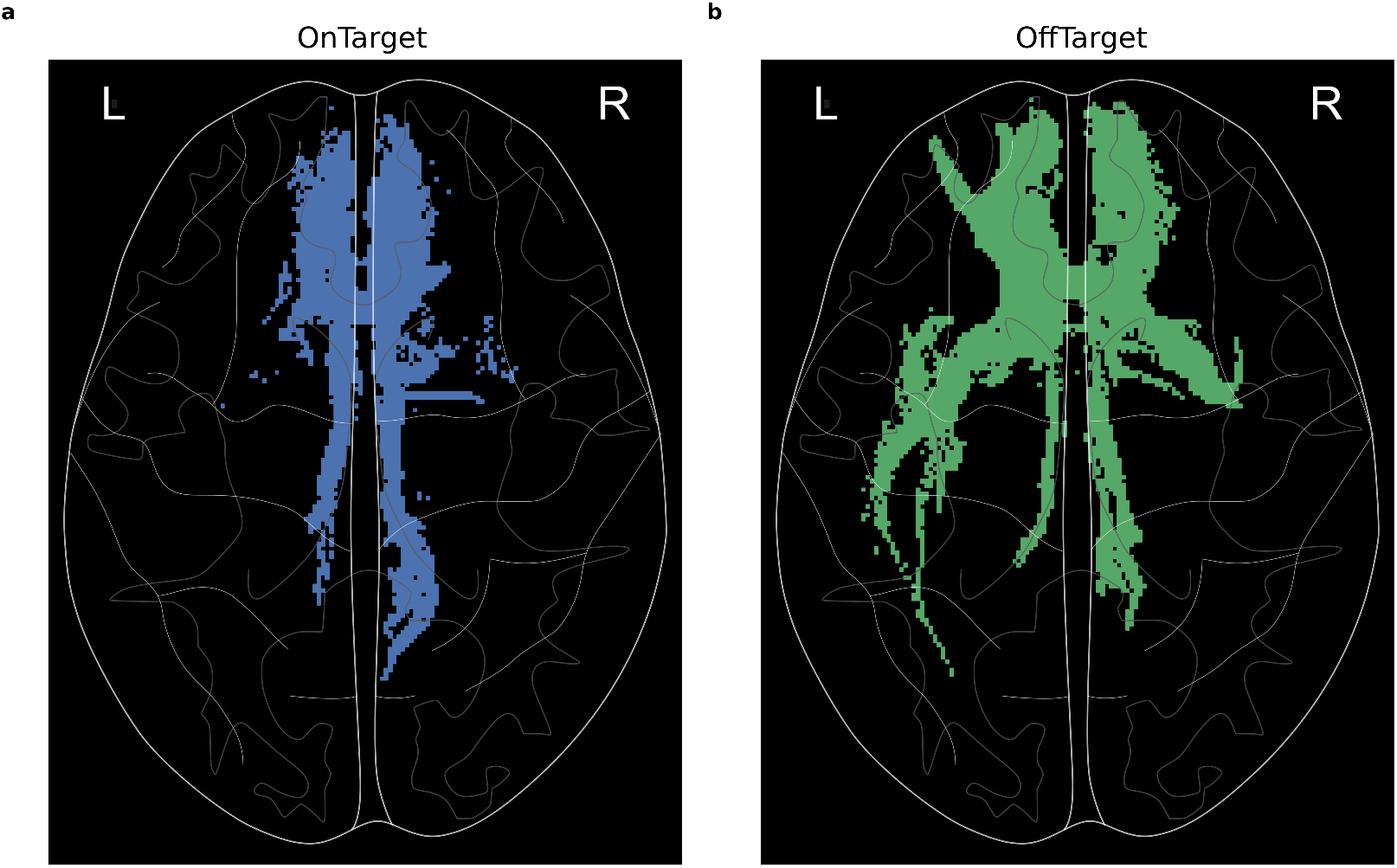
**Engaged tractography at targets** in all patients. **a**, Axial mask of above-threshold voxels in OnTarget vs OffTarget. **b**, Axial mask of above-threshold voxels larger in OffTarget than OnTarget.

## Notes

### Funding Statement

This study was funded by The Whitaker Foundation, National Institutes of Health (UH3NS103550 R01MH106173), Hope for Depression Research Foundation and European Unions Horizon 2020 Framework Programme for Research and Innovation under the Specific Grant Agreement No. 945539 (Human Brain Project SGA3). Implanted devices used in the work were donated by Medtronic, Inc. (Minneapolis, MN).

### Author Declarations

Six consecutive patients were implanted between June 1, 2013 and January 1, 2017 as a part of an IRB approved research protocol at Emory University studying the SCCwm-DBS for TRD (ClinicalTrials.gov Identifier NCT01984710) using inclusion and exclusion criteria described in [1]. Written informed consent was provided by each patient to participate in the study protocol (FDA IDE G130107) and the study was continuously monitored by the Emory University Department of Psychiatry and Behavioral Sciences Data and Safety Monitoring Board.

